## Supplemental appendix 1 for "Convalescent plasma for outpatients with early COVID-19"

Appendix material COMPILE_home_ study

Version, 18^th^ November 2021

### 1 Appendix figures

**Appendix Figure 1.** Identification and inclusion of potential studies

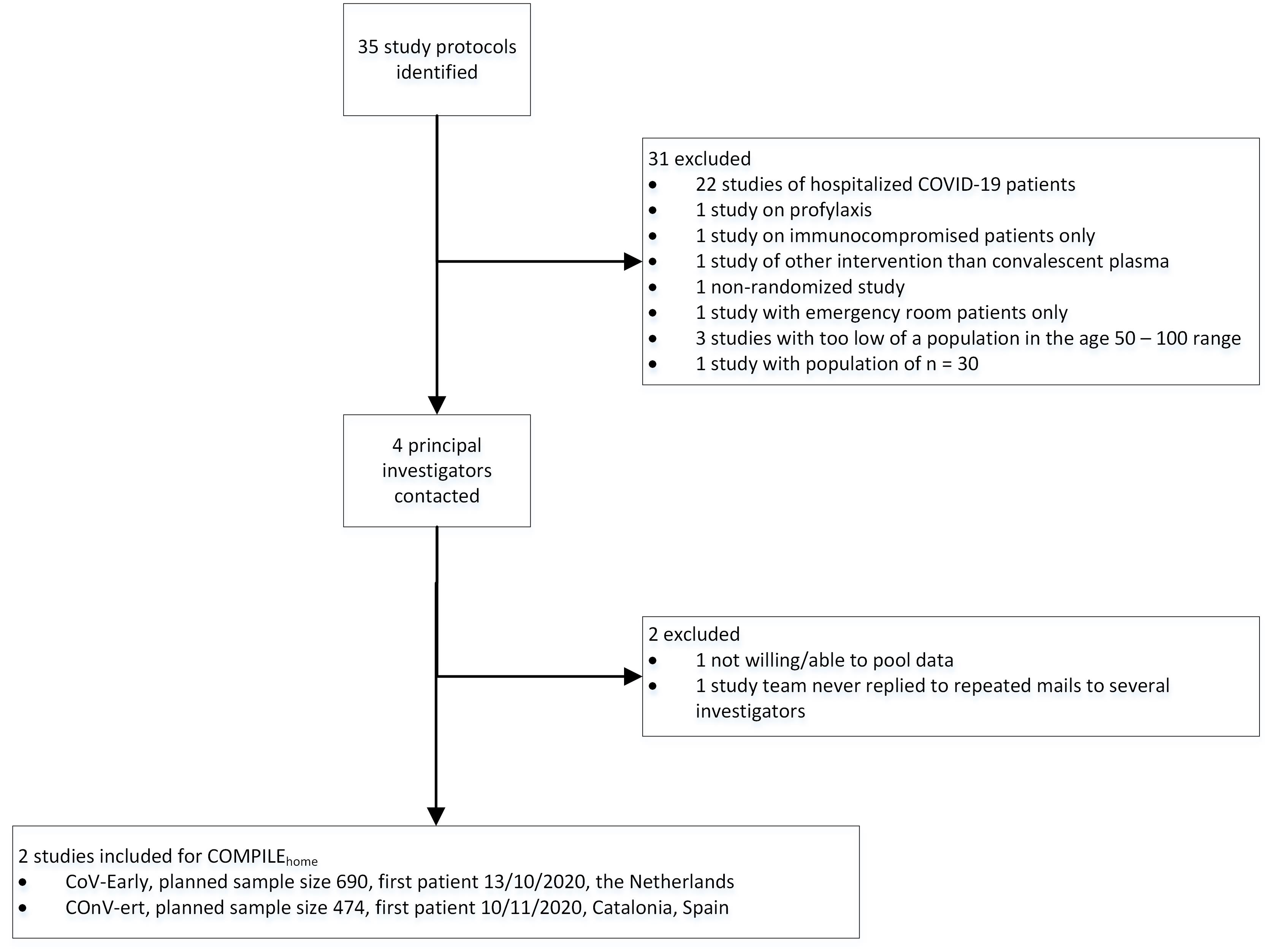

**Appendix Figure 2:** Weekly inclusion rate in CoV-Early and COnV-ert study

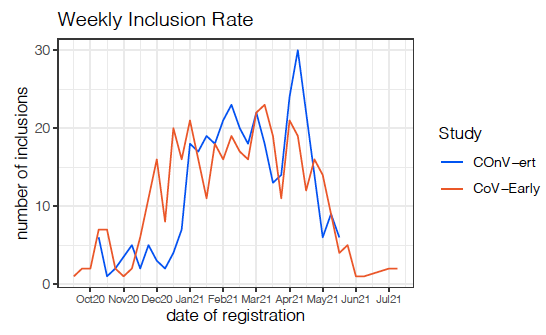

**Appendix Figure 3.** Circulating variants in the Netherlands during recruitment in the CoV-Early study. The first patient was included on 13-10-2020 and recruitment ended 13-07-2021. Available from: https://www.rivm.nl/en/coronavirus-covid-19/virus-sars-cov-2/variants

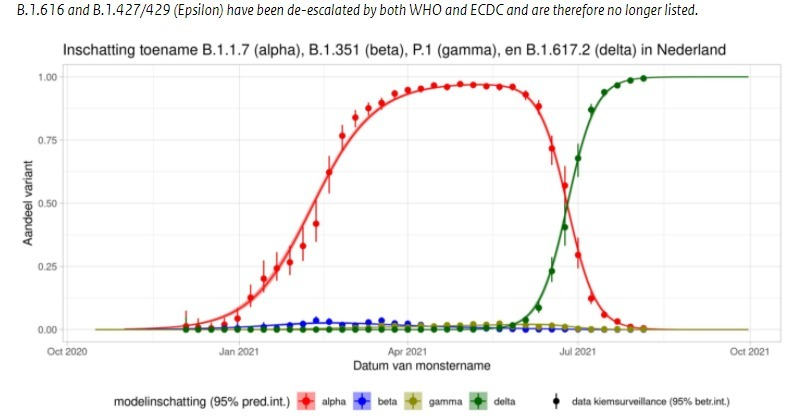

**Appendix Figure 4.** Circulating variants for each week in 2021 in Spain during recruitment in the COnV-ert study. The first patient was recruited on November 10, 2020 and the last patient on May 28, 2021. Note that B.1.351 is the Beta variant (previously known as South-African variant), the B.1.617.2 is the delta variant (previously known as the Indian variant), B.1.1.7 is the alfa or UK variant, while the B.1.621 is also known as the Columbia variant. The grey era (other variants) is assumed to consist almost entirely of the original SARS-CoV-2 virus first isolated in Wuhan. Available from:

https://www.mscbs.gob.es/profesionales/saludPublica/ccayes/alertasActual/nCov/documentos/COVID19_Actualizacion_variantes_20210705.pdf

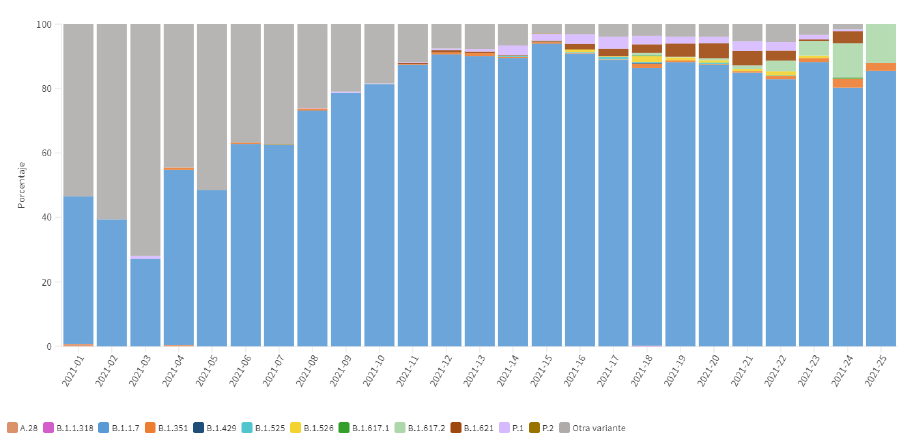

**Appendix Figure 5.** Odds ratios (ORs) of the primary analysis for the 5-point ordinal disease severity scale. Note that an OR <1.0 denotes improved outcome with CP therapy. CP = Convalescent Plasma. TE = Treatment Effect. The OR with 95% credible intervals were 0.936 (0.667-1.311) for treatment with convalescent plasma, 0.827 (0.706-0.970) for a higher oxygen saturation (per standard deviation unit), 0.973 (0.827-1.145) for more days (per standard deviation unit) since COVID-19 symptom onset, 1.170 (0.974-1.404) for each additional comorbidity, 2.615 (1.753-3.896) for inclusion in COnV-ert Trial versus CoV-Early, 1.325 (0.956-1.852) for female sex, 1.033 (0.890-1.198) for age (per year above 50), 0.982 (0.451-2.135) for being immunocompromised. The effect size of convalescent plasma was very comparable for both trials as illustrated by the ORs close to 1 for the difference in TE for both trials.

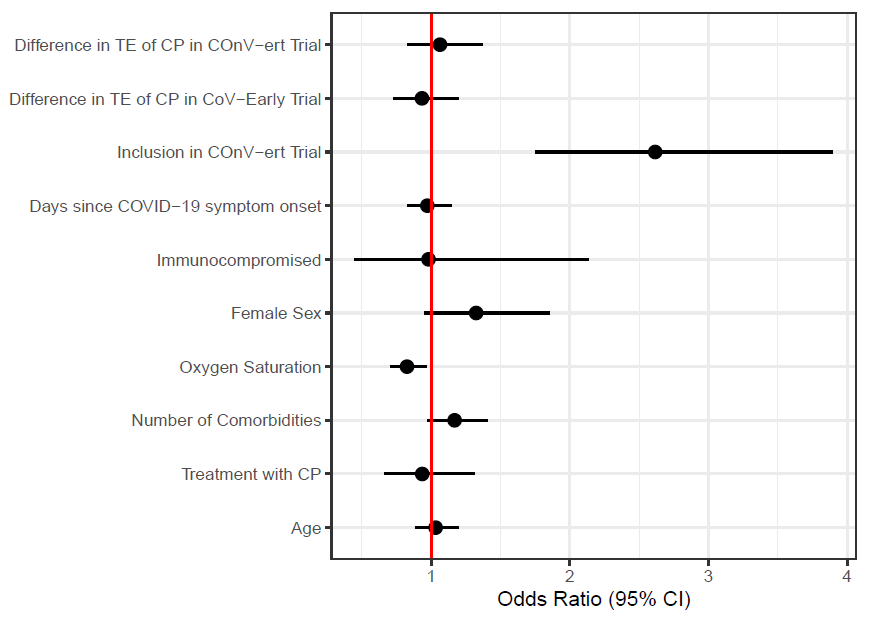

**Appendix Figure 6.** Odds ratio (OR) for improved outcome on the binary endpoint of hospital admission or death. Note that an OR <1.0 denotes improved outcome. CP = Convalescent Plasma. TE = Treatment Effect. The OR with 95% credible intervals were 0.919 (0.592-1.416) for treatment with convalescent plasma, 0.644 (0.508-0.815) for a higher oxygen saturation (per standard deviation unit), (range 91-99%), 1.033 (0.811-1.321) for more days (per standard deviation unit), since COVID-19 symptom onset, 1.178 (0.908-1.507) for each additional comorbidity, 1.679 (0.992-2.810) for inclusion in COnV-ert Trial versus CoV-Early, 1.208 (0.765-1.894) for female sex, 1.033 (0.765-1.266) for age (per year above 50) and 0.773 (0.318-1.896) for being immunocompromised. The effect size of convalescent plasma was very comparable for both trials as illustrated by the ORs close to 1 for the difference in TE for both trials.

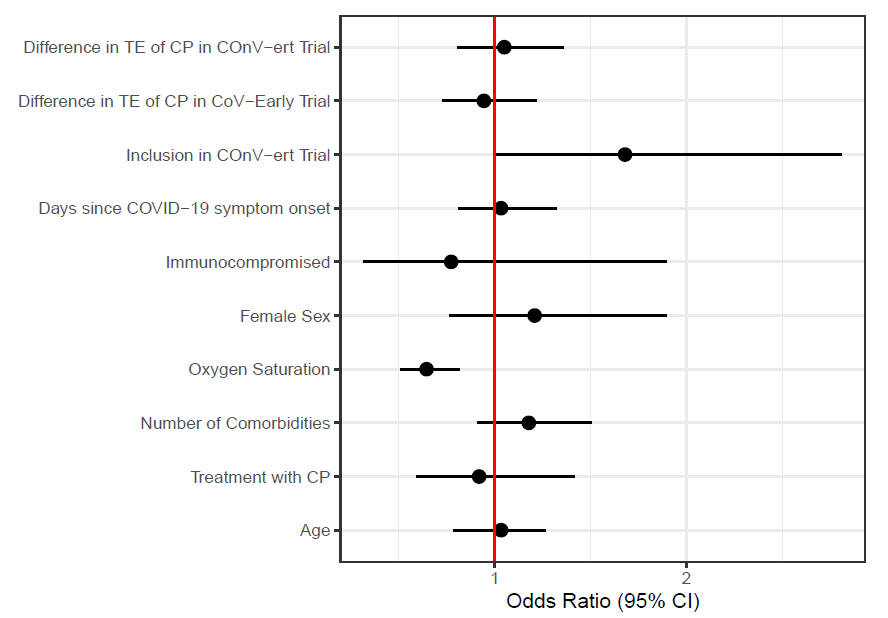

**Appendix Figure 7.** Results of subgroup analysis regarding days since symptom onset and impact of antibody status (positive or negative) of the patient at baseline for the risk of hospital admission or death. OR with 95% confidence intervals are given. Please note that an OR <1.0 denotes improved outcome with CP therapy.

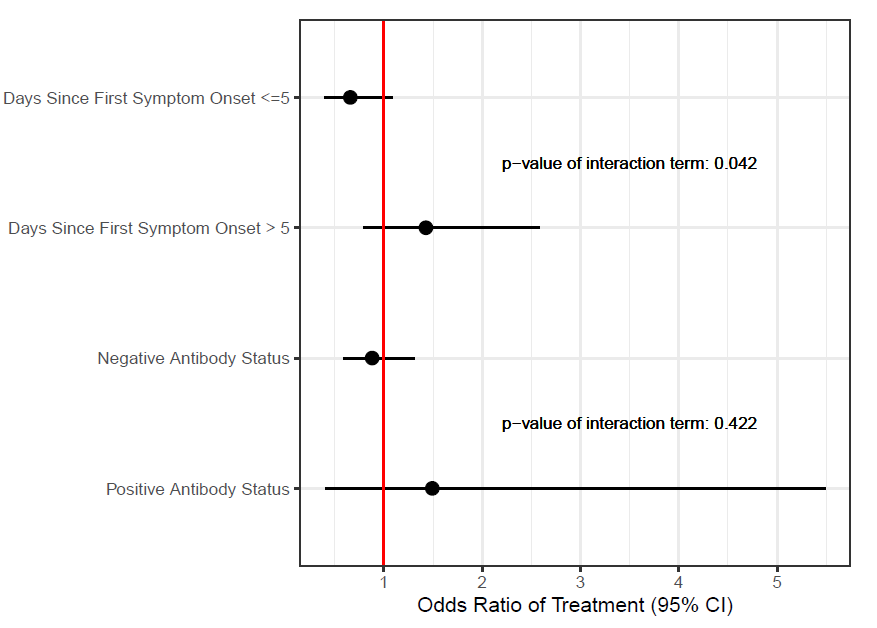

**Appendix Figure 8.** Results of subgroup analysis regarding days since symptom onset, antibody status level at baseline for the 5-point ordinal disease severy scale endpoint. OR with 95% confidence intervals are given. Please note that an OR <1.0 denotes improved outcome with CP therapy.

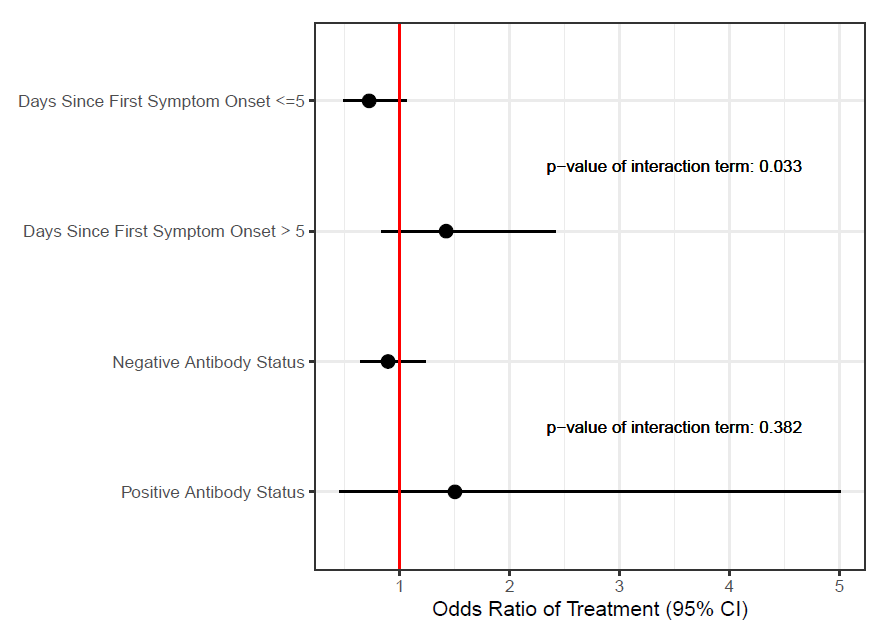

**Appendix Figure 9.** Results of subgroup analysis regarding neutralizing antibody titer level in the convalescent plasma that a patient received for hospital admission or death. OR with 95% confidence intervals are given. Please note that an OR <1.0 denotes improved outcome with CP therapy. The analysis of this secondary endpoint could not be done according to the original statistical analysis plan. This was caused by differences in the way the height of antibody titer was measured in the two studies (continuous versus integer values) as well as the fact that one of both study labs did not dilute serum further once the titer was >1:640 local units (>1:908 IU/mL). To avoid numerical and interpretation issues caused by these differences as well as the low event rates we needed to change the analysis to a dichotomized analysis at using the median titer of 1:386 IU/mL.

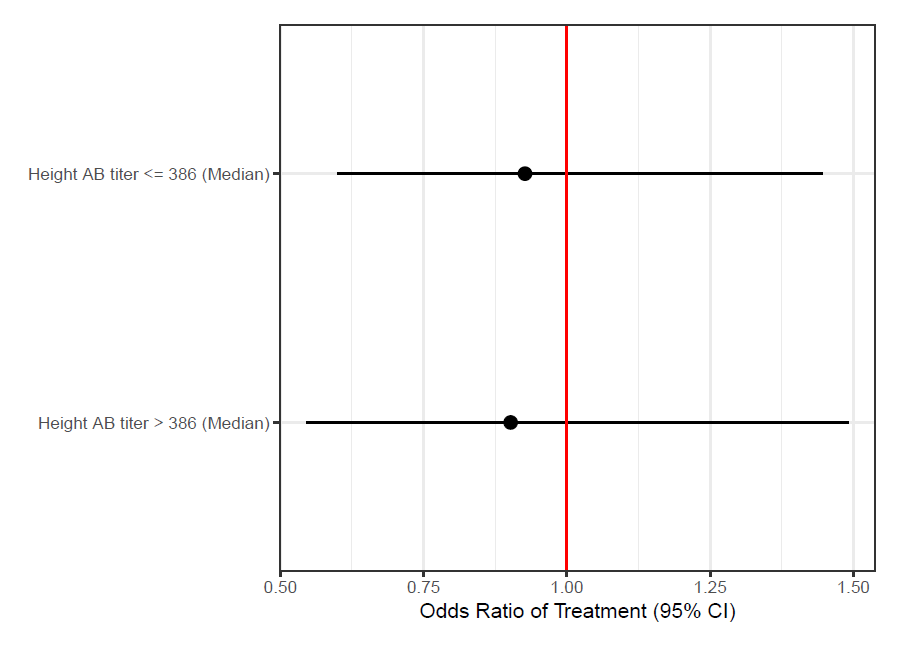

### 2 Appendix tables

**Appendix Table 1.** Comorbidity criteria. Immunodeficiency is not included as a comorbidity. However, it is one of the covariates in the analysis of the primary endpoint and therefore, it is taken into account there. Isolated hypertension without another underlying cardiovascular disease or cardiovascular risk facture is not included as a comorbidity. Abbreviations: AF= atrial fibrillation, BMI = body mass index, CAD= coronary artery disease, COPD = chronic obstructive pulmonary disease; GFR = glomerular filtration rate, HF= heart failure.

|  |  | **Cov-Early** | **COnV-ert** | **Compile_home_** |
| --- | --- | --- | --- | --- |
| **1** | **Obesity** | BMI | BMI | BMI 35 or higher |
| **2** | **Cardiac disease** | AF, HF, CAD, … | HF, CAD | Atrial fibrillation, coronary heart disease, chronic heart Failure, ischemic heart disease, symptomatic atherosclerotic disease other than cardiac |
| **3** | **Lung disease** | COPD or Asthma | COPD and Asthma is registered separately | COPD, Asthma, other chronic lung disease |
| **4** | **Neurological disease** | Stroke or chronic debilitating disease | Cerebrovascular disease | Cerebrovascular disease or chronic debilitating other neurological disease |
| **5** | **Diabetes** | For which medical therapy is given | Any | Diabetes |
| **6** | **Chronic renal failure** | GFR <60 | Is registered but not further defined in CRF | GFR 60 or lower |
| **7** | **Cancer** | Not in complete remission for 1 yr excluding baso/spinocellular skin cancer | Is registered but not further defined in CRF | Cancer not in complete remission for 1 year but excluding baso/spinocellular skin cancer |
| **8** | **Liver disease** | Chronic liver disease leading to cirrhosis or liver dysfunction | Registered in list of other comorbidities | Chronic liver disease with cirrhosis or with liver dysfunction |
| **Other** |  | Not registered | Other significant comorbidities are registered in the list of other comorbidities | Not applicable |

**Appendix Table 2.** Comparison table with similarities and differences between CoV-Early and COnV-ert study protocols. Abbreviations: CKD = chronic kidney disease; eCRF = electronic case report form; IV = intravascular; OD = optical density; PCR = Polymerase Chain Reaction.

|  | **CoV-Early** | **COnV-ert** |
| --- | --- | --- |
| **Inclusion Criteria** | COVID-19, confirmed by PCR or CE-marked antigen test. | Confirmed‌ ‌SARS-CoV-2‌ ‌infection‌ ‌as‌ ‌determined‌ ‌by‌ ‌PCR‌ ‌or‌ ‌validated‌ ‌antigen‌ ‌rapid‌ ‌diagnostic‌ ‌test‌^‌‌^ ‌from‌ ‌nasopharyngeal‌ ‌swabs‌ ‌≤5‌ ‌days‌ ‌prior‌ ‌to‌ ‌inclusion/baseline‌ ‌visit. |
|  | Symptomatic (e.g but not limited to fatigue, fever, cough, dyspnoea, loss of taste or smell, diarrhoea, falls or confusion). | Symptomatic‌ ‌with‌ ‌mild‌ ‌or‌ ‌moderate‌ ‌COVID-19‌ ‌with‌ ‌symptoms‌ ‌onset‌ ‌date‌ ‌≤‌ ‌7‌ ‌days‌ ‌prior‌ ‌to‌ ‌inclusion/baseline‌ ‌visit.‌ ‌ |
|  | 70 years or older OR 50-69 years and 1 or more risk factors OR 18-49 and severely immunocompromised. | Adult‌ ‌male‌ ‌or‌ ‌female‌ ‌individuals‌ ‌of‌ ‌≥50‌ ‌years‌ ‌old.‌ ‌ |
|  |  | Willing‌ ‌to‌ ‌comply‌ ‌with‌ ‌the‌ ‌requirements‌ ‌of‌ ‌the‌ ‌protocol‌ ‌and‌ ‌available‌ ‌for‌ ‌follow-up‌ ‌for‌ ‌the‌ ‌planned‌ ‌duration‌ ‌of‌ ‌the‌ ‌study.‌ |
|  |  | Has‌ ‌understood‌ ‌the‌ ‌information‌ ‌provided‌ ‌and‌ ‌capable‌ ‌of‌ ‌giving‌ ‌informed‌ ‌consent.‌ ‌ |
| **Exclusion criteria** | Life expectancy <28 days in the opinion of the treating physician. |  |
|  | Patient or legal representative is unable to provide written informed consent | Inability‌ ‌to‌ ‌consent‌ ‌and/or‌ ‌comply‌ ‌with‌ ‌study‌ ‌protocol,‌ ‌in‌ ‌the‌ ‌opinion‌ ‌of‌ ‌the‌ ‌investigator.‌ |
|  | Symptomatic for 8 days or more at the time of screening. | (see Inclusion criteria) |
|  | Being admitted to the hospital at the informed consent procedure | Current‌ ‌hospital‌ ‌admission‌ ‌for‌ ‌any‌ ‌cause.‌  Severe‌ ‌or‌ ‌critical‌ ‌COVID-19:‌ ‌  a.Severe‌ ‌COVID-19:‌ ‌respiratory‌ ‌frequency‌ ‌>30‌ ‌breaths‌ ‌per‌  minute,‌ ‌SpO‌_2‌_ ‌<94%‌ ‌on‌ ‌room‌ ‌air‌ ‌at‌ ‌sea‌ ‌level,‌ ‌ratio‌ PaO‌_2‌_/FiO‌_2_ ‌ ‌<300‌ ‌ mmHg,‌ ‌or‌ ‌lung‌ ‌infiltrates‌ ‌>50%.‌ ‌  b. Critical‌ ‌COVID-19:‌ ‌respiratory‌ ‌failure,‌ ‌septic‌ ‌shock,‌ ‌and/or‌ ‌  multiple‌ ‌organ‌ dysfunction.‌ |
|  | Known previous history of transfusion-related acute lung injury- | History‌ ‌of‌ ‌allergic‌ ‌reactions‌ ‌to‌ ‌blood‌ ‌or‌ ‌plasma‌ ‌products‌ ‌or‌ ‌methylene‌ ‌blue.‌  Medical‌ ‌conditions‌ ‌for‌ ‌which‌ ‌2‌0‌‌0‌-300‌ ‌mL‌ ‌of‌ ‌intravenous‌ ‌fluid‌ ‌is‌ ‌considered‌ ‌dangerous‌ ‌(i.e.,‌ ‌decompensated‌ ‌heart‌ ‌failure‌ ‌or‌ ‌renal‌ ‌failure‌ ‌with‌ ‌fluid‌ ‌overload).‌ ‌ |
|  | Known IgA deficiency | Known‌ ‌IgA‌ ‌deficiency‌ ‌with‌ ‌anti-IgA‌ ‌antibodies.‌ ‌ |
|  |  | If‌ ‌female,‌ ‌pregnant‌ ‌or‌ ‌breastfeeding,‌ ‌or‌ ‌planning‌ ‌a‌ ‌pregnancy‌ ‌during‌ ‌the‌ ‌study.‌ ‌ |
|  |  | History‌ ‌of‌ ‌previous‌ ‌confirmed‌ ‌SARS-CoV-2‌ ‌infection.‌ |
|  |  | History‌ ‌of‌ ‌significantly‌ ‌abnormal‌ ‌liver‌ ‌function‌ ‌(Child‌ ‌Pugh‌ ‌C).‌ ‌ |
|  |  | History‌ ‌of‌ CKD‌ ‌‌≥‌‌ ‌stage‌ ‌4,‌ ‌or‌ ‌need‌ ‌of‌ ‌dialysis‌ ‌treatment.‌ ‌ |
|  |  | Any‌ ‌pre-existing‌ ‌condition‌ ‌that‌ ‌increases‌ ‌risk‌ ‌of‌ ‌thrombosis.‌ ‌ |
| **Interventional product** | Single iv infusion of 300 mL of thawed convalescent plasma, preferably ABO-identical  + Standard medical treatment | Single iv infusion of 200-300 mL of convalescent plasma, preferably ABO-identical  + Standard medical treatment |
| **Control arm** | Single iv infusion of 300 mL of thawed non-convalescent plasma (fresh frozen plasma), preferably ABO-identical  + Standard medical treatment | Single iv infusion of 200 to 300 mL of sterile saline solution 0.9%  + Standard medical treatment |
| **Convalescent plasma donor criteria** | Donors had a history of PCR proven symptomatic COVID-19 and had recovered from COVID-19 for at least 14 days. | Donors had a history of PCR proven symptomatic or asymptomatic COVID-19 and had recovered from COVID-19 for at least 28 days. |
|  | Plasma donors were selected based on a virus neutralization titer of at least 1:160. | Plasma donors were selected based on an Euroimmun test with OD of at least 6.0. |
|  | Each donation 600 mL plasma was collected in 2 bags of 300 mL each. Plasma was stored at minus 25 degrees Celsius or colder and tested for pathogens during routine procedures. | Each donation was of a maximum of 600 mL plasma, and it was collected in 2 bags of 200-300 mL each. Plasma was treated with blue methylene and stored at minus 25 degrees Celsius or colder and tested for pathogens during routine procedures. |
|  | Every plasma had a unique identification number by which the product can always be traced back to the donor.  When plasma was administered, this number was registered in the patient file. | Every plasma had a unique identification number by which the product can always be traced back to the donor.  When plasma was administered, this number was registered in the blinded tab of the eCRF for each study participant. |
|  | Administration of conv plasma or fresh frozen plasma was blinded by masking the plasma bag with an opaque bag wrapped around the plasma bag. The transfusion lab personal received the allocation email and wrapped the concealment bag around the plasma bag. | Administration of conv plasma or saline solution was blinded by masking the plasma bag with an opaque bag wrapped around the plasma bag. The transfusion lab received the allocation email. An unblinded study nurse (since the IV perfusion system was not masked) was in charge of the infusion of the iv products. The rest of the investigators and study nurses remained blinded. |

### 3 Appendix methods

The complete CoV-Early and COnV-ert study protocols are available as online supplements. A short summary is given here.

#### COnV-ert study (NCT04621123)

**Trial design**

The COnV-ert study was a multicenter, double-blinded, randomized, controlled trial to assess the efficacy of convalescent plasma in preventing severe COVID-19 in patients infected with SARS-CoV-2 with mild and moderate illness. The trial was conducted between November 2, 2020 and July 28, 2021 at four healthcare centers providing universal healthcare to a catchment population of 3,883,700 in Catalonia, Spain.

The study was conducted according to the Helsinki Declaration of the World Medical Association, and the study protocol was approved by the Ethics Committee at Hospital Germans Trias i Pujol (number PI 20-313) and the institutional review boards of the rest of participating centers. All patients provided written informed consent before enrolling the study, which was supervised by an independent data and safety monitoring board. The trial was registered in ClinicalTrials.gov (NCT04621123).

**Participants**

We included outpatients aged ≥50 years with mild-to-moderate COVID-19 confirmed by RT-PCR or antigen rapid test ≤5 days before randomization and symptoms onset ≤7 days. Mild and moderate COVID-19 were defined according to international guidelines as follows: patients with fever, cough, sore throat, malaise, headache, and muscle pain were considered mild COVID-19, whereas evidence of lower respiratory disease by clinical assessment or imaging and a saturation of oxygen ≥94% on room air was considerate moderate COVID-19. Patients were excluded if they had severe COVID-19 or required hospitalization for any cause, a previous SARS-CoV-2 infection, contraindications with the investigational product, increased thrombotic risk, a history of significantly abnormal liver function (e.g., Child Pugh C), chronic kidney disease stage ≥ 4. Female participants pregnant or breastfeeding or planning a pregnancy during the study were also excluded. Further details on the eligibility criteria are listed in the full protocol that is included as an online supplement.

**Trial Procedures**

Study candidates were identified from two sources: (1) we actively screened the healthcare records of study sites for individuals with evidence of SARS-CoV-2 infection and (2) individuals who tested positive for SARS-CoV-2 infection during epidemiological surveillance could voluntarily register to an institutional website launched by the sponsor and the Catalan Institute of Health. Investigators contacted candidates by phone or in person to inform them about the study, invite them to participate, and check their suitability. Suitable candidates were scheduled a baseline visit, performed either at the hospital or at home by the hospital domiciliary care unit, in which written informed consent was obtained, and the eligibility confirmed.

Eligible patients who provided written informed consent were randomly assigned (1:1) using a computer-generated random-number list to receive one intravenous (IV) infusion of either 200-300 mL of ABO-compatible high-titer convalescent plasma (experimental arm) or 250 mL of sterile saline solution 0.9% (control arm). The study convalescent plasma was selected after being screened for high anti-SARS-CoV-2 IgG titers with ELISA (EUROIMMUN ratio ≥6), according to guidelines, and supplied by the regional blood bank *(Banc de Sang i Teixits de Catalunya* – BST).^1^ Further details on the preparation and characteristics of the plasma are provided in the in the full protocol that is included as an online supplement.

Trained BST staff masked the investigational product with opaque tubular bags that covered the entire infusion catheter to prevent product identification. The masked investigational product was infused over 30 minutes. Patients and all investigators who participated in the trial (including laboratory staff and the statistician) were blinded to treatment allocation, except the baseline study nurses and BST trained personnel, who were not involved in the participants’ follow-up.

Follow-up visits were scheduled on days 7 and 28; additionally, we contacted study patients by phone on days 3, 14, and 60 for assessing their clinical status. During follow-up visits, we obtained blood samples (baseline and day 7) for assessing inflammatory markers and nasopharyngeal swabs (baseline and days 7 and 28) for quantification of SARS-CoV-2 viral load, analyzed by RT-qPCR in a centralized laboratory. Serologic status of all enrolled participants (serum antibody positive or serum antibody negative) were prospectively characterized from baseline samples. All collected data were recorded in an electronic case report form.

**Follow-up**

We defined two primary outcomes regarding treatment efficacy: the clinical outcome was the hospitalization rate on a time frame of 28 days after treatment, and the virological outcome was viral load reduction in nasopharyngeal swabs at day 7 and 28.

Prespecified secondary outcomes were time to complete symptom resolution, change in the 10-point WHO Clinical progression scale score within the 60 days following infusion and change in inflammatory parameters (ferritin, prealbumin, interleukin 6 (IL-6), D-dimer, C reactive protein (CRP)) from baseline to day 7 of follow-up.

Safety was assessed as the proportion of patients with adverse events that occurred or worsened during the follow-up period. Adverse events were assessed for seriousness and causality.

More details are available in the full protocol that is included as on online supplement

#### CoV-Early study (NCT04589949)

**Trial design**

The CoV-Early study was a multicenter, double-blinded, randomized, controlled trial to assess the efficacy of convalescent plasma in preventing severe COVID-19 in patients infected with SARS-CoV-2 with mild and moderate illness at 11 hospitals across the Netherlands. The first patient was included on 13-10-2020 and recruitment ended 13-07-2021.

The study was conducted according to the Helsinki Declaration of the World Medical Association, and the study protocol was approved by the competent authority of the Netherlands (CCMO) and the institutional review board of the Erasmus MC University Medical Center in Rotterdam as well as the board of directors of each of the participating hospitals. All patients provided written informed consent before enrolling the study. The conduct was supervised by an independent data and safety monitoring board. The trial was registered in ClinicalTrials.gov with NCT04589949.

**Participants, recruitment and trial procedures**

Outpatients diagnosed with COVID-19 by PCR or antigen testing and symptomatic for <8 days could be screened. Unless they were severely immunocompromised, they had to be at least 50 years old and have at least one risk factor associated with a higher risk of severe COVID-19. Further details can be found in the full protocol available as an online supplement.

The study was communicated with the Dutch public using all kinds of media including newspapers, medical journals for general practitioners, public health free-of-charge COVID test centers as well as social media. Patients aged 50 or older that tested positive for SARS-CoV-2 at a public health SARS-CoV-2 test centers were contacted by telephone about the positive result of their test by the test center and informed about the possibility of study participation at a nearby hospital. When they showed interest and agreed to be contacted by the study team, their telephone number was shared with the study team and the patient was contacted to get additional information. When the patient fulfilled the in- and exclusion criteria and wanted to participate, he/she received an appointment at the nearest study site the next day. Self-referral was possible as well via [www.cov-early.nl](http://www.cov-early.nl) or [www.coronaplasmastudie.nl](http://www.coronaplasmastudie.nl)

**Screening at study site, baseline visit and follow-up**

All study sites had convalescent plasma for all ABO blood groups available on site. The official screening and baseline visit were done consecutively. Regarding the screening visit, the in- and exclusion criteria were checked again, oxygen saturation was measured to exclude patients with a saturation <93%, written informed consent was obtained, a nasopharyngeal swab was taken, ABO blood group was determined and serum was collected. Eligible patients were randomized using an online randomization tool incorporated in the eCRF (ALEA). The allocation code was mailed to the transfusion lab of the hospital. They provided the study team with one unit of convalescent or control plasma. Masking of investigators and the patient was done by the transfusion lab with the use of a non-transparent concealment bag around the plasma unit. After transfusion, the patient was observed for at least 30 minutes and then could leave the hospital.

**Follow-up**

Patients or when needed their representative or general practitioner were contacted on day 7, day 14 and 28 to evaluate their disease status and severity on a scale from 0 to 5. If a patient had recovered completely (no further symptoms attributable to COVID-19 except for loss of smell or taste), the date of full symptom resolution was registered. Patients that also agreed to participate in a virology and immunology substudy came back to the hospital on several occasions (see full protocol for more details). Patients that participated in the geriatric substudy (age 70 or older) were contacted for a more detailed evaluation (e.g. frailty score, see full protocol for more details).

More details are available in the full protocol that is included as on online supplement

#### Neutralizing antibody testing and inter-laboratory comparison

As both study labs used a different SARS-CoV-2 neutralizing antibody test, a panel of 15 plasma samples was provided for comparison by the Support-E consortium.^2^ This panel included a research reagent 20/130 obtained from the National Institute for Biological Standards and Control (NIBSC, United Kingdom), which had been assigned a unitage of 1,300 international units (IU)/mL of SARS-CoV-2-neutralising antibodies. A further dilution series of a high-titre convalescent plasma sample (initial neutralising antibody titre of 1:5120 provided as neat, and diluted in 1:10, 1:50 and 1:100) was calibrated in IU/mL against this research reagent, and used to assess the linearity of both assays. This allowed retrospective conversion of neutralizing antibody titers into international units (IU/mL) using linear regression formulae derived from assay calibration as shown below for each trials.

**Methods for the neutralizing antibody titer measurement**

**COnV-ert study:** a sample of each convalescent plasma bag was sent to a centralized laboratory (*IrsiCaixa laboratory*) for prospective characterization of neutralizing antibody titers. More than one participant could receive plasma from the same donor. Pseudovirus generation and neutralization assay: HIV reporter pseudoviruses expressing SARS-CoV-2 S protein and Luciferase were generated. pNL4-3.Luc.R-.E- was obtained from the NIH AIDS Reagent Program SARS-CoV-2.^3^ SctΔ19 was generated (GeneArt) from the full protein sequence of SARS-CoV-2 spike with a deletion of the last 19 amino acids in C-terminal, human-codon optimized and inserted into pcDNA3.4-TOPO.^4^ Expi293F cells were transfected using ExpiFectamine293 Reagent (Thermo Fisher Scientific) with pNL4-3.Luc.R-.E- and SARS-CoV-2.SctΔ19 (WH1, B.1.1.7 or B.1.351), at an 8:1 ratio, respectively. Control pseudoviruses were obtained by replacing the S protein expression plasmid with a VSV-G protein expression plasmid as previously reported.^5^ Supernatants were harvested 48 hours after transfection, filtered at 0.45 µm, frozen, and titrated on HEK293T cells overexpressing WT human ACE-2 (Integral Molecular, USA). The neutralization assay has been previously validated in a large subset of samples with a replicative viral inhibition assay.^6^ Briefly, neutralization assays were performed in duplicate in Nunc 96-well cell culture plates (Thermo Fisher Scientific), 200 TCID50 of pseudovirus were preincubated with three-fold serial dilutions (1:60–1:14,580) of heat-inactivated plasma samples for 1 hour at 37ºC. Then, 2x10^4^ HEK293T/hACE2 cells treated with DEAE-Dextran (Sigma-Aldrich) were added. Results were read after 48 hours using the EnSight Multimode Plate Reader and BriteLite Plus Luciferase reagent (PerkinElmer, USA). Neutralization capacity of the plasma samples was calculated by comparing the experimental Relative Light Units (RLU) calculated from infected cells treated with each plasma to the max RLUs (maximal infectivity calculated from untreated infected cells) and min RLUs (minimal infectivity calculated from uninfected cells), and expressed as percent neutralization: %Neutralization = (RLUmax–RLUexperimental)/(RLUmax–RLUmin)*100. The ID50 (reciprocal dilution inhibiting 50% of the infection) was calculated by plotting and fitting the log of plasma dilution vs. normalized response to a 4-parameters equation in Prism 9.0.2 (GraphPad Software, USA).

To facilitate conversion to International Units (IU), a calibrated panel of plasma samples containing a dilution series of a high titer convalescent plasma calibrated in IU/mL using the standard 20/130 obtained from the National Institute for Biological Standards and Control (NIBSC, United Kingdom).^2^ Experimental neutralization titers were converted to IU/mL using the following regression formula (IU/mL = 4160/(2^(Log_2_^(experimentalID50-13.962)/-0.9798^) derived from assay calibration with the pre-quantified control.

**CoV-Early study:** A serum sample from each plasma donor taken on the day of plasma donation was sent to the RIVM lab for virus neutralization testing. Duplicates of two-fold serial dilutions (starting at 1:10) of heat-inactivated sera (30 m, 56°C) were incubated with 100 median tissue culture infectious dose of SARS-CoV-2 strains hCoV-19/Netherlands/ZuidHolland_10004/2020, D614G (WT) and hCoV-19/Netherlands/ NoordHolland_10159/2021 (B.1.351, EVAg, catalog no. 014 V-04058) at 35°C for 1 hour in 96-well plates. Vero E6 cells were added in a concentration of 20,000 cells per well and were incubated for 72 hours at 35°C. The serum virus neutralization titer was defined as the reciprocal value of the sample dilution that showed a 50% protection of virus growth. Samples with titers of ≥20 were defined as SARS-CoV-2 seropositive.

To facilitate conversion to International Units (IU), a calibrated panel of plasma samples containing a dilution series of a high titer convalescent plasma calibrated in IU/mL using the standard 20/130 obtained from the National Institute for Biological Standards and Control (NIBSC, United Kingdom).^2^ Experimental neutralization titers were converted to IU/mL using the following regression formula (IU/mL = 4160/(2^(Log_2_^(experimentalID50-11.832)/-1.146^) derived from assay calibration with the pre-quantified control.
