## Supplementary material for "Convalescent plasma for outpatients with early COVID-19": Reviewers comments nov 2021

Comments for Manuscript "Convalescent plasma for outpatients with early COVID-19. A pooled analysis of two randomized clinical trials."

**Reviewer Comments**

**Reviewer Number:1**

This is an excellent/outstanding paper on a timely topic - namely the early outpatient use of convalescent plasma in high risk COVID-19 positive patients. The key finding is that there was some signal of efficacy in patients treated with 5 days or less of symptom onset.

Other important points include a discussion of the challenges of obtaining truly or exceptionally high titer plasma and the framing of emerging trials showing signals of efficacy for inpatients. That this is a real time "pooling" of data is model for future pandemics and disasters.

A few points to consider. The following papers are perhaps underplayed in terms of the signals of efficacy in the inpatient setting.

Avendaño-Solá et al JCI (N=350) - CP showed a significant benefit in preventing progression to non-invasive ventilation or high-flow oxygen, invasive mechanical ventilation or ECMO, or death at 28 days.

O'Donnell et al JCI (N=223) - 28-day mortality was significantly lower in participants randomized to convalescent plasma versus control plasma (19/150 [12.6%] versus 18/73 [24.6%], OR 0.44, 95% CI 0.22-0.91, p=0.034).

Körper et al JCI (N=101) - A pre-defined subgroup analysis showed a significant benefit for CCP among those who received a larger amount of neutralizing antibodies.

There are also subgroups with efficacy or trends in RECOVERY, REMAPCAP and CONCOR. The later Begin et all paper is the only paper reporting harm, and the harm was only seen with one blood supplier and there was a positive effect of very high titer plasma in late disease seen.

A bit more comparison with the outpatient mAb papers may be helpful in terms of time to treatment. Additionally, a figure or table of the 5 day or less patients would be of interest and/or a KM curve.

The C3PO (Korely)study has a number of issues including a large number of patients in the CP group admitted on the index visit. When these patients are removed from the analysis, CP shows evidence of efficacy.

In summary, this is an excellent paper that fills and important gap. It also paves the way for a series of "whats next" studies and frames a variety of observations that have emerged in the context of the chaos of a pandemic. The "real time" pooling of data is very novel in the context of a pandemic especially.

I enjoyed reading this paper.

**Reviewer Number:2**

A suggested by the title, the manuscript describes a pooled analysis of 2 clinical trials pertaining to use of COVID-19 convalescent plasma in patients with early disease (COVID-19). The negative findings across several clinical outcome measures, largely echo those of studies of moderate to late-stage disease. The methods are clearly described and the limitations are acknowledged appropriately. My comments are mostly minor.

• "The indiscriminate use of poorly selected CP could actually be harmful"; indiscriminate implies that the risk benefit of a given therapy was not considered. That applies to any therapy. There is overwhelming data both from observations studies as well as clinical trials that CCP is safe or no more dangerous that transfusing non-immune (i.e. anti-SARS-CoV-2 negative) plasma in a similar population1-3.

• “However, in this trial patients were recruited at emergency rooms only and were therefore likely to present with more severe symptoms and at a later disease stage.” I might modify the wording to be more explicit about the limitations of the C3PO trial e.g. “However, in this trial patients were recruited at emergency rooms and were therefore more likely to manifest severe symptoms, thus approximating a population with moderate to late-stage disease, contrary to what was intended in the design”.

• In the introduction, one may want to advertise that COMPILE was conceived very early in the CCP initiative, anticipating the downstream challenges pertaining to enrollment and study power.

Methods

• Can the authors please clarify: “Studies were considered suitable when they were RCTs on outpatients recruited within the first 7 days of COVID-19 disease”. On first read, it was unclear as to whether that meant that studies were selected if their inclusion criteria were confined to patients who had symptoms less than 7 days or data were extracted from all RCTs yet for the purposes of the analysis, was confined to those patients who were within 7 days of hospitalization or symptoms onset. This is later clarified in the results but I would state explicitly upfront. I do think that participation of 2 trials out of 35 that ultimately qualified (or even 2 of 4) is a limitation that deserves mention. .

• “data on outpatients with early symptoms are limited” may substitute “rare” for limited

• The authors report important safety findings. I would add some content to the discussion in this regard i.e., the findings are consistent with a host of prior studies that suggest that CCP is safe, with comparable incidence of serious adverse events to that of transfusion of non-immune plasma in a similar patient population.

• The findings from the CCP trials have been largely disappointing (i.e., negative); nonetheless, there have been several large studies and at least 1 clinical trial (cited in this manuscript) that suggest benefit in early disease3-5. Could the authors comment on why there have been such different findings, contributing in part to CCP contentious status.

• Could strengthen your argument about small effect size by stating that CCP’s clinical role is significantly diminished if unable to establish something greater than “a small effect” i.e. it ceases to be practical.

Table 1: baseline characteristics

• Shouldn’t one switch to reporting male —rather than female— sex if the majority were male. It might also make more sense given than male sex was a risk factor for severe COVID-19

• Low numbers of comorbidities: could there still be a role for CCP in selected patient subsets?

3. Arnold Egloff SA, Junglen A, Restivo JSA, Wongskhaluang M, Martin C, Doshi P, Schlauch D, Fromell G, Sears LE, Correll M, Burris HA, LeMaistre CF. Convalescent plasma associates with reduced mortality and improved clinical trajectory in patients hospitalized with COVID-19. The Journal of Clinical Investigation 2021.

4. Libster R, Pérez Marc G, Wappner D, Coviello S, Bianchi A, Braem V, Esteban I, Caballero MT, Wood C, Berrueta M, Rondan A, Lescano G, Cruz P, Ritou Y, Fernández Viña V, Álvarez Paggi D, Esperante S, Ferreti A, Ofman G, Ciganda Á, Rodriguez R, Lantos J, Valentini R, Itcovici N, Hintze A, Oyarvide ML, Etchegaray C, Neira A, Name I, Alfonso J, López Castelo R, Caruso G, Rapelius S, Alvez F, Etchenique F, Dimase F, Alvarez D, Aranda SS, Sánchez Yanotti C, De Luca J, Jares Baglivo S, Laudanno S, Nowogrodzki F, Larrea R, Silveyra M, Leberzstein G, Debonis A, Molinos J, González M, Perez E, Kreplak N, Pastor Argüello S, Gibbons L, Althabe F, Bergel E, Polack FP. Early High-Titer Plasma Therapy to Prevent Severe Covid-19 in Older Adults. N Engl J Med 2021;384: 610-8.

5. Joyner MJ, Carter RE, Senefeld JW, Klassen SA, Mills JR, Johnson PW, Theel ES, Wiggins CC, Bruno KA, Klompas AM, Lesser ER, Kunze KL, Sexton MA, Diaz Soto JC, Baker SE, Shepherd JRA, van Helmond N, Verdun NC, Marks P, van Buskirk CM, Winters JL, Stubbs JR, Rea RF, Hodge DO, Herasevich V, Whelan ER, Clayburn AJ, Larson KF, Ripoll JG, Andersen KJ, Buras MR, Vogt MNP, Dennis JJ, Regimbal RJ, Bauer PR, Blair JE, Paneth NS, Fairweather D, Wright RS, Casadevall A. Convalescent Plasma Antibody Levels and the Risk of Death from Covid-19. New England Journal of Medicine 2021;384: 1015-27.

**Reviewer Number:3**

The authors discuss the early administration of convalescent plasma (CP) in high-risk patients with Covid-19. This manuscript delineates the clinical outcomes after therapeutic intervention with CP. The topic has become very controversial over the last months, with literature supporting both standpoints. Overall, the paper is well written and represents important new information concerning the use of CP in the ongoing pandemic; the authors are to be commended for this piece. My comments are minor:

Title:
- The title is descriptive and reflective of the manuscript's content.

Methods - Study patients and selection criteria:

-> "Although the exact inclusion and exclusion criteria could vary across the trials, all the subjects had to fulfil the following criteria; 1) Participant of a trial that joined the COMPILEhome consortium 2) Confirmed COVID-19 diagnosis by a diagnostic PCR or antigen test."

- PCR results are likely to be an insensitive method because even in a patient who has fully recovered from COVID, PCR can remain positive for weeks or even longer.

Methods - Intervention:
- Additional detail is needed to describe how the titer was determined.

Results:
-> "74 patients were hospitalized, 6 required mechanical ventilation, and 3 died."

- More information on these patients should be provided.

- Discussion:
-> "In this analysis of 782 patients with COVID-19 randomized to high-titer CP or placebo within 7 days of disease onset"

-Please elaborate on the reason for selecting this 1-week period instead of a shorter time frame. An additional 2-3 sentences elaborating on this matter may be helpful here. Moreover, it is important to note that ± 70% of the patients will have passed the 7d symptoms window when they are admitted to the hospital. Different patients will have varying degrees of disease course. Some elderly people only show typical COVID-19 symptoms until later stages of the disease, and some may have asymptomatic infection.

-> "It is the largest of its kind, studying the effect of CP for high-risk outpatients with COVID-19 early after initiation of symptoms."

-Kindly define "high-risk outpatients". Moreover, the low median age of 58 years is not representative of a high-risk population.

**Reviewer Number:4**

The work brings together two trials with similar characteristics in different countries, which in a scenario where the delta variant was low (5%), its results are comparable with studies such as C3PO. It is very solid and I think the authors provide very useful information to help understand the role of convalescent plasma in patients with 7 or less days of symptoms onset. It is likely that studies of these characteristics can no longer be carried out with a growing vaccinated population.
